# TM6SF2 Determines Both the Degree of Lipidation and the Number of VLDL Particles Secreted by the Liver

**DOI:** 10.1101/2023.06.23.23291823

**Authors:** Gissette Reyes-Soffer, Jing Liu, Tiffany Thomas, Anastasiya Matveyenko, Heather Seid, Rajasekhar Ramakrishnan, Steve Holleran, Norann Zaghloul, Carole Sztalryd-Woodle, Toni Pollin, Henry N. Ginsberg

## Abstract

In 2014, exome-wide studies identified a glutamine176lysine (p.E167K) substitution in a protein of unknown function named transmembrane 6 superfamily member 2 (TM6SF2). The p.E167K variant was associated with increased hepatic fat content and reduced levels of plasma TG and LDL cholesterol. Over the next several years, additional studies defined the role of TM6SF2, which resides in the ER and the ER-Golgi interface, in the lipidation of nascent VLDL to generate mature, more TG-rich VLDL. Consistent results from cells and rodents indicated that the secretion of TG was reduced in the p.E167K variant or when hepatic TM6SF2 was deleted. However, data for secretion of APOB was inconsistent, either reduced or increased secretion was observed. A recent study of people homozygous for the variant demonstrated reduced in vivo secretion of large, TG-rich VLDL1 into plasma; both TG and APOB secretion were reduced. Here we present new results demonstrating increased secretion of VLDL APOB with no change in TG secretion in p.E167K homozygous individuals from the Lancaster Amish community compared to their wild-type siblings. Our in vivo kinetic tracer results are supported by in vitro experiments in HepG2 and McA cells with knock-down or Crispr-deletions of TM6SF2, respectively. We offer a model to potentially explain all of the prior data and our new results.

## Introduction

During the past several decades, epidemiologic and genetic evidence have indicated that, in addition to the plasma concentration of cholesterol-rich low density lipoproteins (LDL), levels of plasma triglyceride-rich lipoproteins (TGRL) and their remnants are also causally related to risk for ASCVD ^1^. Increased levels of circulating TGRL result mainly from increased rates of assembly and secretion of very low density lipoproteins (VLDL). The latter is a large macromolecular (30-90 nm diameter) aggregate consisting of more than 20,000 molecules of neutral core-lipids (TG and cholesterol-esters) with a coating of several thousand molecules of amphipathic phospholipids and free cholesterol, along with one molecule of apolipoprotein B (APOB100) and a small number of other apolipoproteins ^2^. VLDL is not only the precursor of atherogenic apolipoprotein B100-lipoproteins, including TGRL-remnants, intermediate density lipoprotein (IDL), and LDL, but also plays a critical role in the maintenance of hepatic TG content. As might be expected, the assembly and secretion of VLDL is complex, with over 40 proteins presently identified as participating in a multi-step process that includes the translocation and initial lipidation of APOB100 to generate a nascent VLDL; subsequent addition of lipid, mainly TG in the ER, to form a mature VLDL; complex vesicular transport of VLDL to the Golgi for further addition of lipids as well as glycosylation of APOB100; and finally, the secretion of VLDL from hepatocytes ^3–5^.

Based on the importance of TG transport from the liver via VLDL secretion, it was to be expected that the search for genes associated with altered hepatic lipid homeostasis, particularly non-alcoholic fatty liver disease (NAFLD), would identify several that are intimately involved in the lipidation of APOB100. The critical importance of APOB100 and microsomal triglyceride transfer protein (MTP) in VLDL assembly and maintenance of normal levels of hepatic TG has been known for several decades, but the list of additional proteins critical to this process has increased significantly in the past 20 years^6^. A major breakthrough in the search for genes involved in steatosis came in 2008 with the identification of an isoleucine148methionine substitution in patatin-like phospholipase domain-containing protein 3 (PNPLA3) in a genome-wide association study (GWAS) of the Dallas Heart Study Cohort ^7^. Although PNPLA3 is not involved in VLDL assembly and secretion, its discovery energized the field and led to the identification of a number of additional NAFLD-associated genes, including reports of a group of variants in a locus on chromosome 19 that included more than a dozen genes^8, 9^. Those studies culminated in simultaneous reports in 2014 of exome-wide studies identified a glutamine176lysine (p.E167K) substitution in a protein of unknown function named transmembrane 6 superfamily member 2 (TM6SF2) ^10, 11^. The p.E167K variant was associated with increased hepatic fat content and reduced levels of plasma TG and LDL cholesterol.

In the past 10 years, several studies in rodents and cultured hepatocytes have demonstrated a role for TM6SF2 in the lipidation of nascent VLDL. The protein has been localized to the ER and pre-Golgi (where lipidation of nascent VLDL occurs) in human hepatoma lines ^10^ and in mouse and rat primary hepatocytes ^12, 13^. However, physiologic studies, including knockdown, knockout, or over-expression of TM6SF2 in cells and rodents have produced variable results regarding the development of steatosis and the secretion of TG and APOB100. Although a detailed discussion of those data is beyond the scope of this paper, the discrepancies suggested to us that in vivo tracer-kinetic studies conducted in subjects homozygous for the p.E167K substitution (referred subsequently as KK, compared to their unaffected EE siblings), complemented by hepatoma-based cell studies, could provide additional insights relevant to the differences in the literature that remained after many earlier studies. While we were carrying out our studies, Boren and colleagues published results from their *in vivo* tracer kinetics in unrelated EE and KK individuals ^14^. Our results differ significantly from those of Boren et al, adding further uncertainty to the field. We believe, however, that the differences in the two human kinetic studies, together with previous data from cells and rodents, and our new cell data, allow us to offer a potentially unifying model of how TM6SF2 affects the assembly and secretion of VLDL from the liver.

## Methods for Human Studies

### Study Subjects

We recruited five participants homozygous for the p.E167K variant in TM6SF2, hereafter denoted by KK, and five gender-matched, unaffected siblings, hereafter denoted by EE, ages 25 to 65 years, from the Lancaster Old Order Amish population ^15^. Participants were not receiving lipid altering medications. All study participants provided written informed consent and the studies were approved by the Institutional Review Boards of the University of Maryland School of Medicine and the Columbia University Irving Medical Center (CUIMC).

### Stable Isotope Kinetic Studies

The two sibs in each pair were studied on the same day at the Amish Research Clinic (ARC) in Lancaster, PA. The protocol for these studies was one that we have used previously ^16^, with minor modifications to allow subjects to have some samples obtained at their homes. On day 1, participants fasted for 12 hours, after which a nurse visited their homes and drew baseline bloods for safety, and fasting lipid and lipoprotein measurements. After 6pm on Day 1, they were NPO until 1am on day 2 when they started a liquid, isocaloric, 18% fat diet that was provided every two hours for the next 30 hours. At 5:30am (Day 2), they arrived at the ARC, where two IV’s were placed in antecubital veins of each arm and baseline bloods were drawn (time 0hr). Immediately after, boluses of ²H₃-L-leucine (10 µmol/kg BW), Ring-¹³C₆-L-phenylalanine (29.4 µmol/kg BW), and ²H₅-glycerol (100 µmol/kg BW) were administered over a 10-minute period, followed by a constant infusion of ²H₃-L-leucine (10 µmol/kg BW per hour) over 15 hours. Additional blood samples were collected at 20 and 40 minutes, and at 1, 2, 4, 6, 8, 10, 12, 14, 15, 15.2, 15.4, 16 hours after the administration of tracers and processed to isolate plasma and serum. Following the16hr blood sample, the subjects returned to their homes where they continued to consume the liquid meals. Eight hours later a nurse drew the final 24hr blood sample in their homes. VLDL, IDL, LDL, and high density lipoprotein (HDL) were obtained from the 16 plasma samples that were collected at the ARC and shipped to CUIMC where they were processed by sequential density ultracentrifugation ^17^.

### Determination of stable isotope enrichment of APOB100 and TG

The isolated lipoprotein fractions were used to determine stable isotopic enrichments of ²H₃-L-leucine and Ring-¹³C₆-L-phenylalanine in APOB100 in VLDL, IDL, and LDL. ApoB100 was isolated from VLDL, IDL, and LDL by SDS-polyacrylamide gel electrophoresis. The isolated APOB100 bands were excised from the gels, hydrolyzed, and the amino acids derivatized. Plasma free amino acids were recovered from 0.25 mL plasma after precipitation of proteins with acetone and extraction of the aqueous phase with hexane. The aqueous phase was dried under vacuum, amino acids were derivatized. Enrichments of -^2^H_3_-leucine and Ring-^13^C_6_-phenylalanine tracers in APOB100-lipoproteins and plasma free leucine and phenylalanine were measured by gas chromatography - mass spectrometer (GCMS) using an Agilent 6890 gas chromatography and a 5973 mass spectrometer with negative chemical ionization. Additionally, kinetic analysis of TG in VLDL was performed with ²H₅-glycerol. TG was separated from phospholipid by zeolite binding. The TG was resolubilized with chloroform. We performed trans-esterification with methanolic HCL. Glycerol was isolated by liquid/liquid extraction (hexane and water added) and derivatized to triacetin (glycerol tri-acetate) through incubation with acetic anhydride. We performed GC/MS positive chemical ionization (CI) with selective ion monitoring (SIM) of m/z 159 and 164.

### Compartmental modeling of APOB100 and TG metabolism

Fractional clearance rates (FCR) and production rates (PR) of TG and APOB100 in VLDL, and of APOB100 in IDL and LDL were determined using a compartmental model to fit stable isotope enrichment data ^17–19^. In our general model, APOB100 and TG are required to have the same pool structure and the same rate constants for each VLDL pool, but with different mass distributions. With more than one pool in the VLDL fraction, the different mass distributions lead to different VLDL FCRs for TG and APOB, since VLDL FCR is obtained as a weighted average of the individual FCRs (the weights given by the mass distribution). However, if there is only one pool in the VLDL fraction, TG and APOB necessarily have the same FCR, which was the case here. For each study, the minimum number of pools needed to simultaneously fit the nine sets of data (two tracers, leucine and phenylalanine, in VLDL-, IDL-, and LDL-APOB100 and plasma amino acids, and one tracer, glycerol, in VLDL-TG) was chosen for the final model. In the present study, the final model had one pool each for VLDL, IDL, and LDL. The data were fitted by least squares, giving equal weight to all data points (that is, assuming a constant error variance for all measurements) using a computer program, Poolfit ^18^, which solves the differential equations in closed form and computes the fits and parameter sensitivities as sums of exponentials. The fits yielded fractional catabolic rates (FCRs) of APOB100 in VLDL, IDL, and LDL, and TG in VLDL. The model also estimated rates of conversion of APOB100 between VLDL, IDL, and LDL. Production rates (PRs in mg/kg/day) were calculated by multiplying FCRs (in pools/day) by the appropriate lipoprotein pool sizes of APOB100, which were estimated as each lipoprotein’s concentration of APOB100 in mg/ml multiplied by the plasma volume (45 ml/kg). Based on the best fit of the present data, we have only one VLDL pool for APOB and TG, conversion from VLDL to LDL via IDL, no direct out pathway from IDL, and a small component of direct secretion of LDL from the liver.

### Biochemical and immunologic assays

Day 1 bloods were collected after a 12-hr overnight fasting period. Additional timed blood samples were collected while the subjects were consuming the liquid diet, both before (0hr) and at various time points after the stable isotope infusion was started (20min, 40min, 1, 2, 4, 6, 8, 10, 12, 14, 15, 15.2, 15.4, 16, 24 hrs.). Plasma cholesterol, TG and HDL-cholesterol (C) were measured by Integra400plus (Roche). Plasma LDL-C levels were estimated using the Friedewald formula. Cholesterol and TG were also measured enzymatically in VLDL, IDL, LDL, and HDL isolated by ultracentrifugation. Plasma APOC2, APOC3 and APOE were measured by human enzyme-linked immunosorbent assay (ELISA) kits (ab168549 [APOC2]; ab154131 [APOC3]; ab108813 [APOE], Abcam, Cambridge, MA). ApoB100 in plasma and in VLDL, IDL, and LDL was measured using an APOB100 ELISA kit (A70102 AlerCheck, Inc.).

### Statistical analyses for human studies

Paired t-tests were used to compare variables between sibling-pairs. Significance for major endpoints of plasma and lipoprotein levels of APOB, VLDL APOB100 PR and FCR, and VLDL TG PR and FCR, were set at p<0.05. Significance for all other endpoints was set at p<0.01.

## Methods for Cell Studies

### Cell culture protocols

Human hepatoma cell line HepG2 and a rat hepatoma cell line McA-RH7777 (McA) were both purchased from ATCC (Rockville, USA). HepG2 cells were maintained in Dulbecco’s modified Eagle’s medium (DMEM) supplemented with 0.1 mM nonessential amino acids, 1 mM sodium pyruvate, penicillin (100 U/ml), streptomycin (100 U/ml), and 10% fetal bovine serum. McA cells were grown in DMEM supplemented with 10% fetal bovine serum,10% horse serum, penicillin (100 U/ml) and streptomycin (100 U/ml), in a 5% CO2 incubator at 37°C.

### Knockdown or overexpression of TM6SF2 by lentivirus in HepG2 cells

Lentiviral hRNAs targeting *TM6SF2* plasmids were obtained from Dharmacon (GE healthcare) with the following sequences: pGIPZ-shRNA1: CTTTGTTCCTCTTGCACCA; pGIPZ-shRNA2:AAAGACACCCGGAGATGTC; pGIPZ-shRNA3:ACATCAGCATCTGCACCTT. HEK 293T cells were co-transfected with lentiviral vector and the packaging plasmids pMD2.G (AddGene) and psPAX2 (AddGene). The cell culture medium containing the viral particles was collected after 48 and 72 hours of culture, and viral particles were concentrated by ultracentrifugation. Then HepG2 cells were transduced with shRNA lentiviruses or control non-silencing lentivirus using 8 μg/ml polybrene at an MOI of 4 according to the manufacturer’s instruction. The cells were harvested at 24 or 48 h for isolation of total RNA or protein extract to verify the efficiency of *TM6SF2* knockdown.

To construct the lentiviral plasmid carrying TM6SF2, human TM6SF2 cDNA was acquired by PCR amplification with BamHI and Nhel cutting sites from the OmicsLink vector (EX-EGFP-M02, genecopoeia) and cloned into the pLOCTurboRFP vector (Dharmacon, GE Healthcare). The constructs containing TM6SF2 cDNA was confirmed with Sanger sequencing. To generate the lentiviruses overexpressing TM6SF2, we used the same protocol as described above.

### Tm6sf2 gene knockout by CRISPR/Cas9

The sgRNAs targeting rat *Tm6sf2* genes (NM_001127654) at exon 1 or exon 3 were designed using the CRISPR design websites. The three target sequences were 5′TGGTTGAGTACGTAGGACAC-3′ (T1), 5′-CGAAACTTGGTTGAGTACGT-3′ (T2), and 5′-ATGGACATCCCGCCGCTAGC-3′ (T3). The sgRNAs were synthesized and cloned into plentiCRISPR v2 (Addgene) according to Sanana et al. ^20^ and the constructs are followed by sequencing validation (Genewiz). To produce the lentiviruses containing sgRNA and Cas9, the tm6sf2-sgRNA plasmids were co-transfected with the packaging plasmids pVSVg (AddGene) and psPAX2 (AddGene) into HEK293T cells. After 72 h, cell supernatants containing viruses were collected. To generate *Tm6sf2* knockout cells, McA cells were transduced with titrated virus using 8 μg/ml polybrene, and selected in media using 1μg/ml puromycin. The cells were isolated by <u>serial dilution</u> and went through an expansion period to get clonal cell lines. Finally, two stably *Tm6sf2* knockout cell clones were obtained at a multiplicity of infection of 0.5. Genomic DNA was extracted using QuickExtract DNA Extraction Solution (Epicenter) from control and knockout clonal cells. The genomic region about 650 bp was PCR amplified with high-fidelity Herculase II Fusion DNA polymerases (Agilent Technologies). The PCR primers were forward 5-′TGGGTGCAGGTAAG GGGCGTTA-3′ and reverse 5′-TGCCAGGGGAGTTGGGTGAGTT-3′, and the PCR products were separated in 1% agarose gel and purified with a gel extraction kit (Thermo Fisher Scientific) for Sanger DNA sequencing. Knockout efficiency was determined using the ICE analysis from Synthego.

### Measurement of the synthesis and secretion of APOB

We used the method of Pan et al. ^21^ to label APOB100 and APOB48 for steady state or pulse chase studies with/without oleic acid (OA) in HepG2 or Mca cells. The cells were pre-incubated with methionine (Met)-free medium (Invitrogen) with or without 0.4 mM OA for 2 hrs before the media were replaced by Met-free medium with [^35^S]-Met (150uCi, PerkinElmer) and incubated for 2 hrs or for 20 min with or without 0.4 mM OA. In some experiments, cells were chased for various times with new, unlabeled media and cells and media were harvested for immunoprecipitation of APOB100 and APOB48 with a specific antibody (MilliporeSigma). Samples were resolved by 3.5% SDS-PAGE with the quantity loaded based on the trichloroacetic acid–perceptible radioactivity in cell lysates or media. The gel was treated with Autofluor (National Diagnostic), dried, and exposed to x-ray film (Fisher Scientific) at –70 °C.

### Measurement of the synthesis and secretion of radiolabeled TG

McA cells were labeled with [^14^C] OA (10uCi/ml) for 6hrs and washed with PBS three times. Lipids from the cells were extracted with 2 ml of 3:2 n-hexane:isopropanol (v/v) at room temperature for 2 hrs. The extraction solution was collected and dried N_2_ gas. The media were extracted with mixing 20 volumes of chloroform/methanol (2/1, v/v) and 5 volumes of H_2_O, and spun for 5 min at 2,000 rpm. The lower organic phase was collected into glass tubes and dried under N2. Lipid extracts were spotted on a TLC Silica gel 60 plastic sheet (MiliporeSigma). Lipids were separated by thin-layer chromatography employing a mixture of 70:30:1 hexane:diethyl ether:acetic acid (v/v) as a mobile phase followed by lipid visualization in an iodine chamber. The separated TG was scraped and analyzed by liquid scintillation counting.

### Quantification of triglyceride levels in McA cells

TG content of McA cells was measured using triglyceride assay Kit (Abcam 65336) according to the manufacturer’s instructions. 1.5 x 10^7^ cells were washed with cold PBS three times and homogenized in 1ml of 5% NP-40 solution. homogenized samples were heated to 80 – 100°C in a water bath until the NP-40 solution becomes cloudy, and insoluble material was removed using a microcentrifuge. All samples were diluted 10-fold with ddH2O before proceeding. 2 ul lipase was added into each well containing 50 ul of each standard, sample, and or background to convert triglyceride to glycerol and fatty acid. After 20 min, 50 ul of the reaction mixture was added into each sample well in the plate. Then the plate was incubated at room temperature for 60 minutes, and absorption was measured on a microplate reader at OD 570 nm. A calibration curve of a standard solution was used to

### Western blot analysis

The level of endoplasmic reticulum stress was examined by western blotting for proteins involved in the uncoupled protein response, Cultured cells were washed twice with PBS, and lysed using a lysis buffer containing 25 mM Tris-HCl, 2 mM Na_3_VO_4_, 10 mM NaF, 10 mM Na_4_P_2_O_7_, 1 mM EGTA, 1 mM EDTA, 1% NP-40, 5 μg/ml leupeptin, 5 μg/ml aprotinin, 10 nM okadaic acid, 1 mM PMSF and phosphatase inhibitors (Millipore). Protein extracts were separated on a 4-12% SDS-PAGE gel (Bio-rad) and transferred to 0.45-μm nitrocellulose membrane(Bio-rad). Membranes were incubated overnight at 4 °C with primary antibody, and the protein bands were detected with HRP-conjugated secondary antibodies and SuperSignal West Pico enhanced chemiluminescent solution (Pierce). The primary antibodies used were anti–phospho-eIF2α (Cell Signaling Technolog), eIF2α (Cell Signaling Technology), IRE1a (Cell Signaling Technology), BIP (Cell Signaling Technology), Chop (Cell Signaling Technology) and monoclonal anti–β-actin (Sigma-Aldrich).

### BODIPY staining for lipid droplets

Cells were grown at a density of 0.5 × 10^6^ cells/well on 6-well plates containing coverslips (Fisher Scientific) pre-coated with collagen (Sigma-Aldrich). The cells were washed with PBS three time, incubated with 0.4mM oleic acid (Sigma) for 4h. A neutral lipid-specific BODIPY (boron-dipyrromethene 4,4-difluoro1,3,5,7,8-pentamethyl-4-bora-3a,4a-diaza-s-indacene 493/503; Molecular Probes) was diluted in PBS at a concentration of 1 mg/mL and applied to cells for 30 min at 37°C. Then the cells were washed twice and fixed with 4% paraformaldehyde (Thermo Fisher Scientific) for 30 min at room temperature. Following fixation, the samples were washed three times and mounted with ProLong Gold Antifade reagent (Thermo Fisher Scientific) with 4,6-diamidino-2-phenylindole (DAPI, Thermo Fisher Scientific). Images were acquired using a Leica TCS SP8 confocal microscope, equipped with a 63 × 1.4 lens and diode and argon lasers.

### VLDL particle size by negative staining electron microscopy

VLDL in the medium was separated by sucrose gradient ultracentrifugation as described above. A total of 1ml of VLDL fraction from the top of tube was collected. To use electron microscopy to image VLDL particles, the VLDL fraction were fixed in 2% paraformaldehyde in PBS, and kept at 4°C. 5 μl of fixed suspension was added onto glow discharged carbon coated 400 mesh Cu/Rh grid (Ted Pella Inc., Redding, CA), and stained with 1% aqueous uranyl acetate (Polysciences, Inc, Warrington, PA). Stained grids were imaged under Talos120C transmission electron microscope (Thermo Fisher Scientific, Hillsboro, OR) using Gatan OneView digital camera (4K x 4K, Gatan, Inc., Pleasanton, CA). The size distribution of the particles was determined using ImageJ software (10 random images/group).

### Next-Generation sequencing and data analysis

Total RNA was isolated from *Tm6sf2* knockout cells or control cells using a Qiagen RNA extraction kit (74104). Poly-A pull-down was used to enrich mRNAs from total RNA samples, then RNA-seq libraries were constructed using Illumina TruSeq RNA sample Prep Kit (Illumina, San Diego, CA, USA) following the manufacturer’s instruction. Libraries are then sequenced on Illumina NovaSeq 6000 at Columbia GenomeCenter. There were multiplex samples in each lane, which yields targeted numbers of paired-end 100bp reads for each sample. We use RTA (Illumina) for base calling and bcl2fastq2 (version 2.19) for converting BCL to fastq format, coupled with adaptor trimming. We perform a pseudoalignment to a kallisto index created from transcriptomes (Rat: GRCr38) using kallisto (0.44.0). We test for differentially expressed genes under various conditions using Sleuth or DESeq2, R packages designed to test differential expression between two conditions. A heatmap was generated using transcripts per million (tpm) data. A Z-score normalization was calculated for the normalized tpm values for each transcript by subtracting the mean and then dividing by the standard deviation. The computed Z score was then used to plot a heatmap by R studio. Genes with dark red colors are up-regulated and those with blue colors are down-regulated.

### Statistical analyses for cell experiments

All statistical analyses (identified in the figure legends) were performed with GraphPad Prism 5.0. Significance for any differences between groups was set at 0.05.

## Results of human studies

### Study Population

We enrolled 5 gender and aged matched (within 10 years) sibling-pairs, each pair having one affected KK and one unaffected EE individual. There was one male pair and 4 female pairs. Mean ages (KK 45.6±11.3, EE 41.8±10.8), BMIs (KK 28.5±9,3 EE 25.8.0±3.2), and fasting plasma glucose levels (KK 88.6±7/1, EE 83.8±92) ere similar between the groups (**Table 1**).

**Table 1.** Baseline Data.

|  | **Affected KK** | **Unaffected EE** | **Difference** | **paired t**<br>**p-value** |
| --- | --- | --- | --- | --- |
| **Sex** | 1M/4F | 1M/4F |  |  |
| **Age (years)** | 45.6±11.3 | 41.8±10.8 | 3.8±5.9 | 0.23 |
| **BMI kg/m<sup>2</sup>** | 28.5±9.3 | 25.8±3.2 | 2.6±6.6 | 0.42 |
| **Glucose mg/dL** | 88.6±7.1 | 83.8±9.2 | 4.8±7.5 | 0.22 |
| **Total Cholesterol mg/dL** | 141.8±19.7 | 152.8±27 | -10.9±16.7 | 0.22 |
| **HDL Cholesterol mg/dL** | 53.2±16.4 | 50.2±13.6 | 3±8.4 | 0.47 |
| **Triglycerides mg/dL** | 97.6±56 | 102.5±55.7 | -4.9±26.2 | 0.70 |
| **LDL Cholesterol mg/dL** | 69±11.3 | 82.3±25.1 | -13.3±19.6 | 0.20 |
| **APOCII µg/mL** | 37±18.3 | 49.6±25.7 | -12.6±13 | 0.10 |
| **APOCIII µg/mL** | 101.2±25.4 | 110±32.3 | -8.8±26.9 | 0.51 |
| **APOE µg/mL** | 130.7±46.5 | 126.2±65.7 | 4.4±29 | 0.75 |

All biochemical values were obtained on Day 1 of the study after a 12 hour fast as defined in Methods. HDL – high density lipoprotein; LDL- low density lipoprotein; APO- apolipoprotein. BMI – body mass index (weight in kilograms divided by height in meters squared).

### Plasma Lipid and Apolipoprotein Levels

Plasma total cholesterol, HDL-C, TG, and LDL-C were not different between KK and EE siblings (**Table 1**). Plasma levels of APOC2, APOC3, and APOE were also not different between the two groups.

### Lipoprotein APOB100 and lipid levels

There were no differences between KK and EE sibling-pairs for levels of plasma, VLDL-, IDL-, or LDL-APOB100 concentrations (**Table 2**). Plasma levels of APOB48 were lower in KK versus EE individuals (p=0.03) but VLDL APOB48 levels were similar in the two groups. There were no differences between sibling pairs for cholesterol or TG levels in the isolated lipoprotein fractions (**Table 2**).

**Table 2.** Lipoprotein Lipids and Apolipoproteins.

|  | **Affected KK** | **Unaffected EE** | **Difference** | **paired t**<br>**p-value** |
| --- | --- | --- | --- | --- |
| **Plasma APOB100 mg/dL** | 65.4±11.9 | 75.9±4.1 | -10.6±11.7 | 0.11 |
| **VLDL-APOB100 mg/dL** | 5.1±3 | 5.2±2.9 | -0.1±2.4 | 0.90 |
| **IDL-APOB100 mg/dL** | 0.9±0.7 | 1.5±1 | -0.6±1.6 | 0.45 |
| **LDL-APOB100 mg/dL** | 40.2±9 | 44.4±4.3 | -4.2±6.8 | 0.24 |
| **Plasma APOB48 mg/dL** | 1.4±0.9 | 1.6±0.9 | -0.3±0.2 | 0.03 |
| **VLDL-APOB48 mg/dL** | 0.38±0.38 | 0.41±0.41 | -0.03±0.01 | 0.48 |
| **VLDL-Chol mg/dL** | 13.8±9.9 | 16.2±11.2 | -2.4±5.1 | 0.35 |
| **VLDL-TG mg/dL** | 66.9±51.4 | 78.3±58.2 | -11.4±16.9 | 0.21 |
| **IDL-Chol mg/dL** | 3.9±1.3 | 4.3±2.4 | -0.5±2.2 | 0.65 |
| **IDL- TG mg/dL** | 5.3±1.6 | 6±2.5 | -0.7±1.9 | 0.47 |
| **LDL-Chol mg/dL** | 64.1±7.1 | 71.4±18.9 | -7.4±13.5 | 0.29 |
| **LDL-TG mg/dL** | 11.4±4 | 13±5 | -1.7±3 | 0.28 |
| **HDL-Chol mg/dL** | 40.1±9.5 | 37±7.7 | 3.1±5.7 | 0.29 |
| **HDL-TG mg/dL** | 11.5±4.8 | 10.3±3.5 | 1.2±4.2 | 0.55 |

All levels are the group means of individual means of 3 samples obtained from each participant during the 24 hour sampling period for the kinetic studies. Apo – apolipoprotein; VLDL – very low density lipoprotein; IDL- intermediate density lipoprotein; LDL- low density lipoprotein; HDL- high density lipoprotein. Chol- cholesterol; TG- triglyceride.

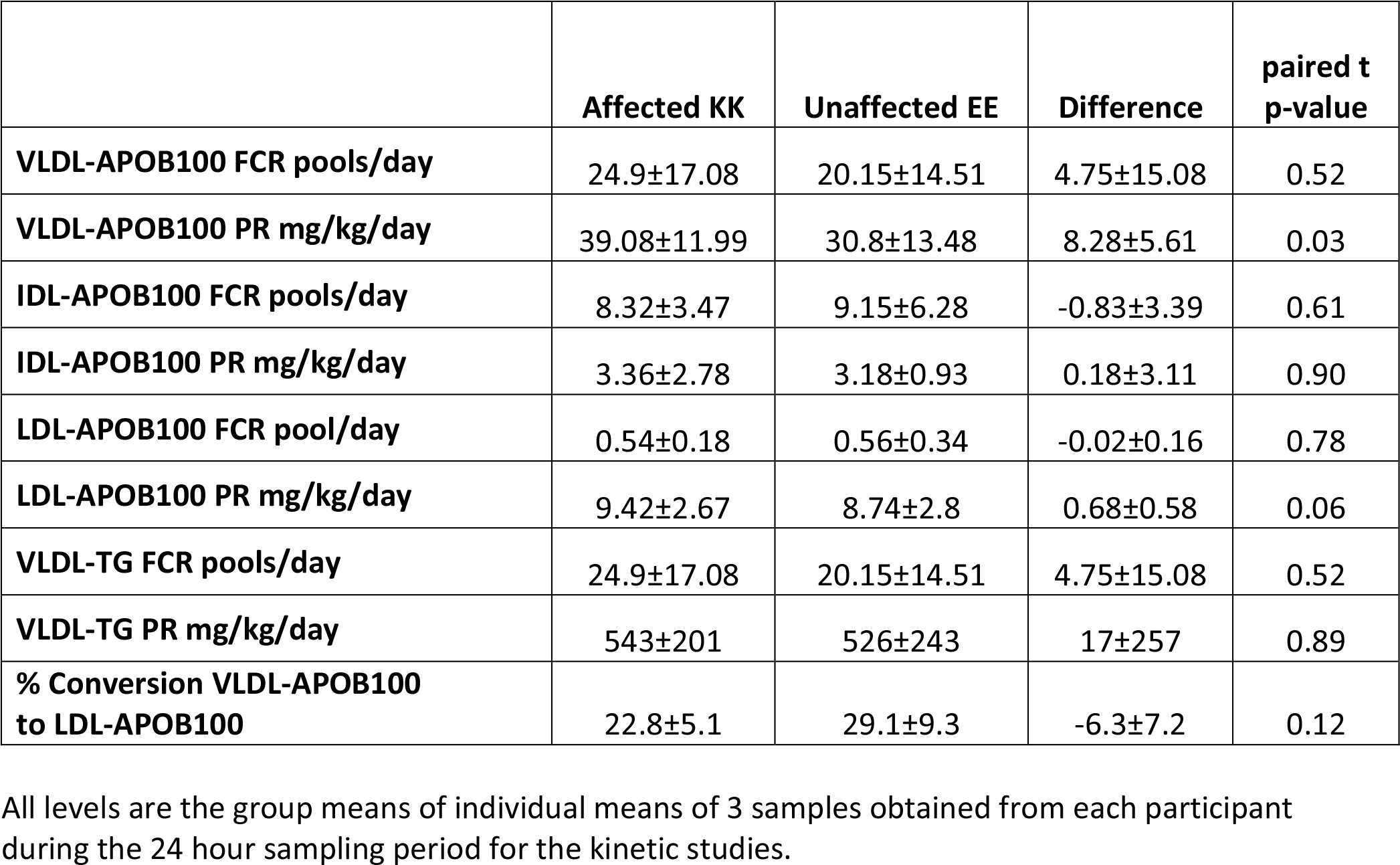
TM6SF2 (KK) Individuals Have Increased VLDL-ApoB100 Production Rates Compared to (EE) Siblings.

|  | **Affected KK** | **Unaffected EE** | **Difference** | **paired t**<br>**p-value** |
| --- | --- | --- | --- | --- |
| **VLDL-APOB100 FCR pools/day** | 24.9±17.08 | 20.15±14.51 | 4.75±15.08 | 0.52 |
| **VLDL-APOB100 PR mg/kg/day** | 39.08±11.99 | 30.8±13.48 | 8.28±5.61 | 0.03 |
| **IDL-APOB100 FCR pools/day** | 8.32±3.47 | 9.15±6.28 | -0.83±3.39 | 0.61 |
| **IDL-APOB100 PR mg/kg/day** | 3.36±2.78 | 3.18±0.93 | 0.18±3.11 | 0.90 |
| **LDL-APOB100 FCR pool/day** | 0.54±0.18 | 0.56±0.34 | -0.02±0.16 | 0.78 |
| **LDL-APOB100 PR mg/kg/day** | 9.42±2.67 | 8.74±2.8 | 0.68±0.58 | 0.06 |
| **VLDL-TG FCR pools/day** | 24.9±17.08 | 20.15±14.51 | 4.75±15.08 | 0.52 |
| **VLDL-TG PR mg/kg/day** | 543±201 | 526±243 | 17±257 | 0.89 |
| **% Conversion VLDL-APOB100**<br>**to LDL-APOB100** | 22.8±5.1 | 29.1±9.3 | -6.3±7.2 | 0.12 |

All levels are the group means of individual means of 3 samples obtained from each participant during the 24 hour sampling period for the kinetic studies.

### APOB100 and TG Metabolism

As noted in Methods, we modeled VLDL-TG and VLDL-APOB100 metabolism jointly, and the best fits of the stable isotope enrichment data required only a single VLDL pool. Thus, the FCRs for VLDL-APOB100 and VLDL-TG in each subject were the same. There were no differences in the FCRs for VLDL-, IDL-, or LDL-APOB100 or the FCR for VLDL-TG between the KK and EE individuals (**Table 3**). However, the PRs of VLDL-APOB100 were 39.1±12 and 30.8±14 mg/kg/day in the KK and EE groups, respectively, a difference of 38.3±47.5% that was statistically significant (p=0.03). In contrast, there was no difference in the VLDL-TG PR between the two groups (543±201 and 526±243; p=0.88). The ratio of VLDL-TG PR to VLDL-APOB100 PR, an indicator of the size of newly secreted VLDL particles, were 18.3±7.7 for EE and 14.7±6.9 for KK (p=0.07). Thus, the mean size of newly secreted VLDL particles was smaller in the KK group, consistent with reduced lipidation of each VLDL particle.

The PRs for IDL-APOB100 were similar in the KK and EE siblings (3.36±2.78 and 3.18±0.93 mg/kg/day; p=0.90). There was a strong trend for a greater LDL-APOB100 PR in the KK versus the EE groups (9.42±2.67 and 8.74±2.8; p=0.06). The numerically greater LDL-AP0B100 PR in the KK individuals was consistent with their greater VLDL-APOB100 PR together with the similar rates of conversion of VLDL-APOB100 to LDL-APOB100 (**Table 3**). Two of the 10 studies required a small direct input of LDL APOB100 from the liver (data not shown).

## Results of cell studies

### Knockdown of TM6SF2 gene expression in HepG2 cells

The human hepatoma cell-line, HepG2, was transduced with either a Lente virus carrying an shRNA targeting *TM6SF2* or a scrambled shRNA and examined for expression of *TM6SF2* by qRT-PCR (**Figure 1A** upper panel) or TM6SF2 protein by Western blotting (**Figure 1A** lower panel). Two lines were identified that had very significant reductions in expression and one line was chosen for further experiments. When cells transduced with either *TM6SF2* shRNA (KD) or scrambled shRNA (Ctrl) were labeled with [35S] methionine for 2-hrs, there were no differences in newly synthesized, radiolabeled APOB100 in the cells (**Figure 1B**). There was, however, significantly greater secretion of newly synthesized, radiolabeled APOB100 into the media from the KD cells. Next, the effects of oleic acid (OA) on APOB100 secretion were examined in the same HepG2 cell-lines, Cells were incubated with or without 0.4 mM OA for 2-hr and then labeled with [35S] methionine for 20 min and chased for 60 min with or without OA (**Figure 1C**) Addition of OA resulted in similar, modest increases in newly synthesized, radiolabeled APOB100 in both the CT (white bars) and KD (dark bars) in both cells (left panel) and media (right panel).

**Figure 1A.**
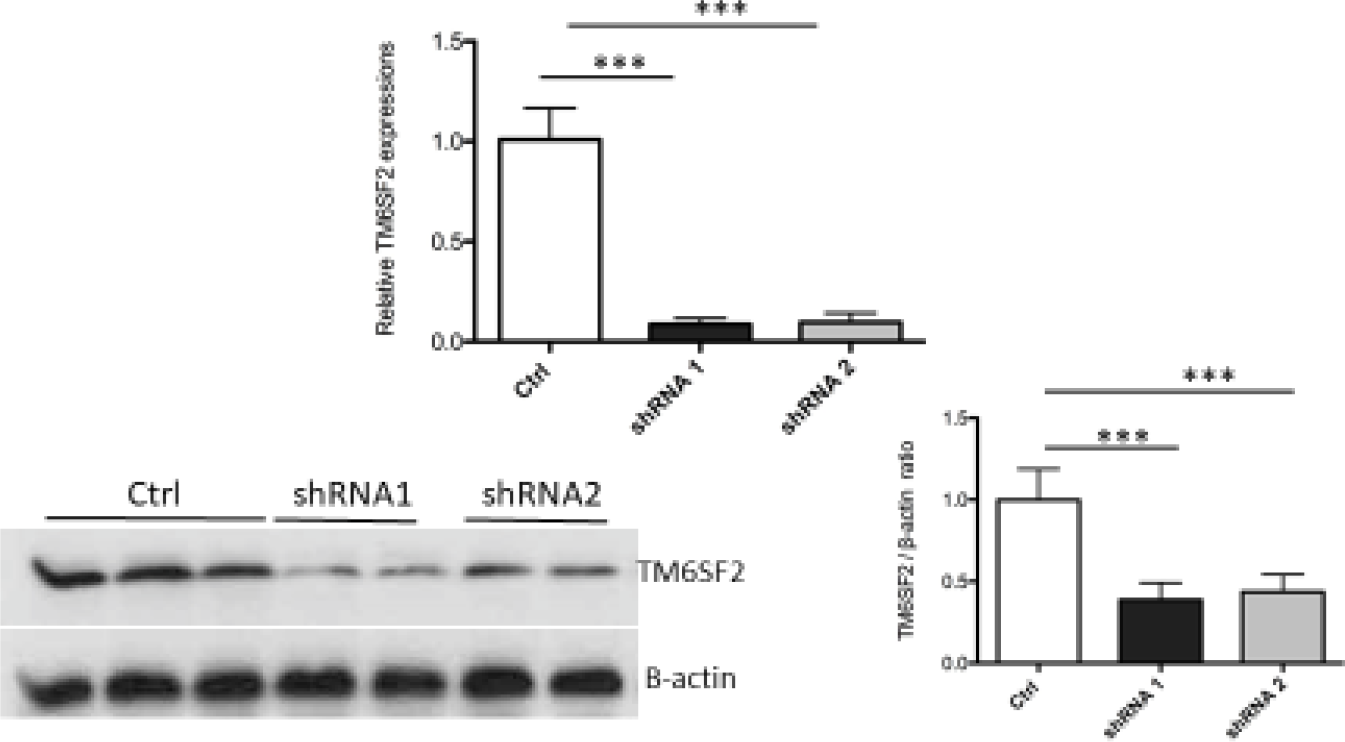
*TM6SF2* silencing was confirmed by qRT-PCR and western blotting. HepG2 cells were transduced with shRNA targeting *TM6SF2* (shRNA 1 and 2) or scramble shRNA (Ctrl) using lentiviral particles. Upper panel: qRT-PCR for TM6SF2 gene expression from cells expressing shRNA1, shRNA2, and scrambled shRNA. Lower panel (left): TM6SF2 expression levels by Western blotting in two TM6SF2 shRNA knockdown cells and Ctrl cells. Lower panel (right): Densitometric analysis of TM6SF2 protein normalized by B-actin. Each bar represents the data from 2 experiments respectively. ***p < 0.001.

**Figure 1B.**
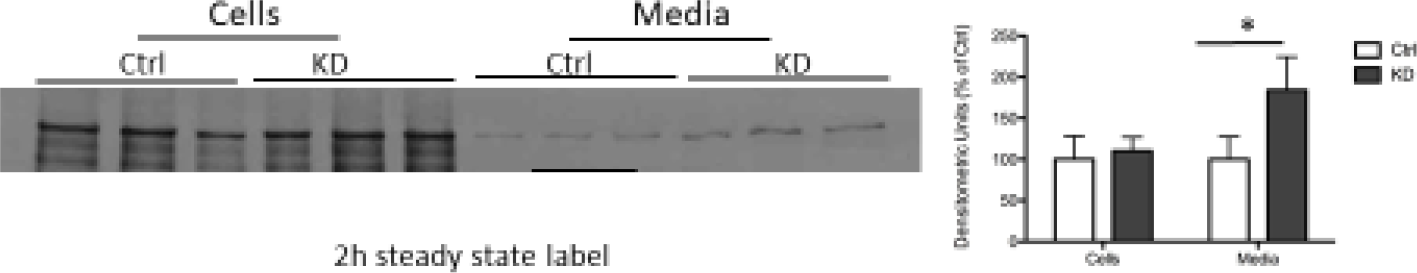
APOB100 secretion with steady state labeling. Upper: The cells were labeled with [^35^S] methionine for 2 hours. Then the cells and media were harvested and ApoB100 was immunoprecipitated prior to SDS PAGE and autoradiography. The data are expressed the mean ± standard deviation of three independent experiments. *p < 0.05; ns: not significant.

**Figure 1C.**
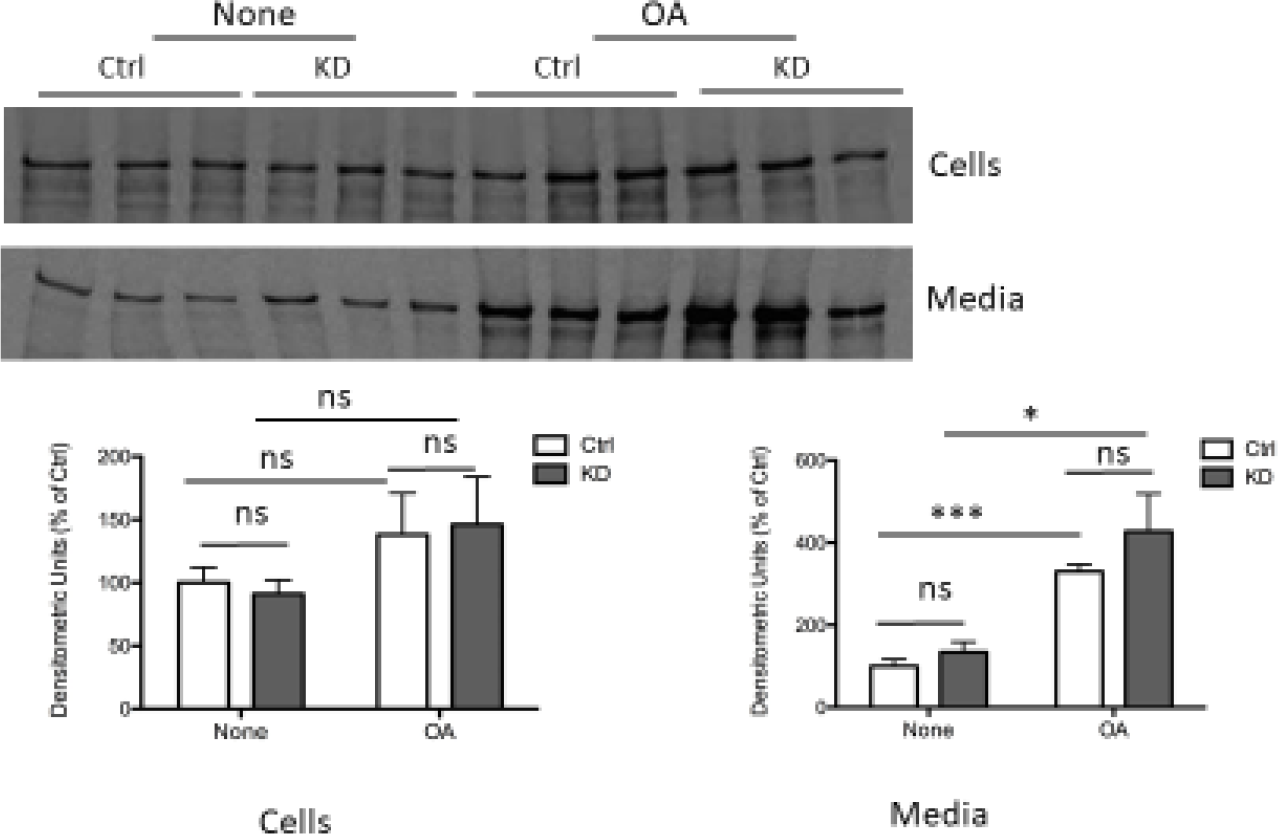
ApoB secretion in TM6SF2 knockdown cells treated with oleic acid (OA). The cells were pre-incubated with 0 4mM OA for 2h, then labeled with [^35^S] methionine for 20min following 60 min chase. Then the cells and media were harvested and ApoB was immunoprecipitated prior to SDS PAGE and autoradiography. The data are expressed the mean ± standard deviation of three independent experiments, ns: not significant. *p<0 05, **·p < 0.001; ns: not significant.

### Overexpression of TM6SF2 in HepG2 cells

*TM6SF2* overexpression in HepG2 cells was achieved as described in Methods. **Figure 2A** shows the relative fold-expression of WT (EE) *TM6SF2* estimated by qRT-PCR compared to expression in cells receiving empty plasmid (Mock) (**top**) and the qualitative increases in protein by Western blotting (**bottom**). Overexpression of WT (EE) TM6SF2 resulted in significantly reduced levels of newly synthesized, radiolabeled APOB100 in cells and nearly complete loss of secretion of APOB100 into the media compared to cells receiving empty plasmid (Mock) (**Figure 2B**). The marked reduction of cell and media APOB100 in cells overexpressing TM6SF2 was confirmed in a pulse-chase experiment where cells were labeled for 20 min and chased in unlabeled media for 2 hrs (**Figure 2C**).

**Figure 2A.**
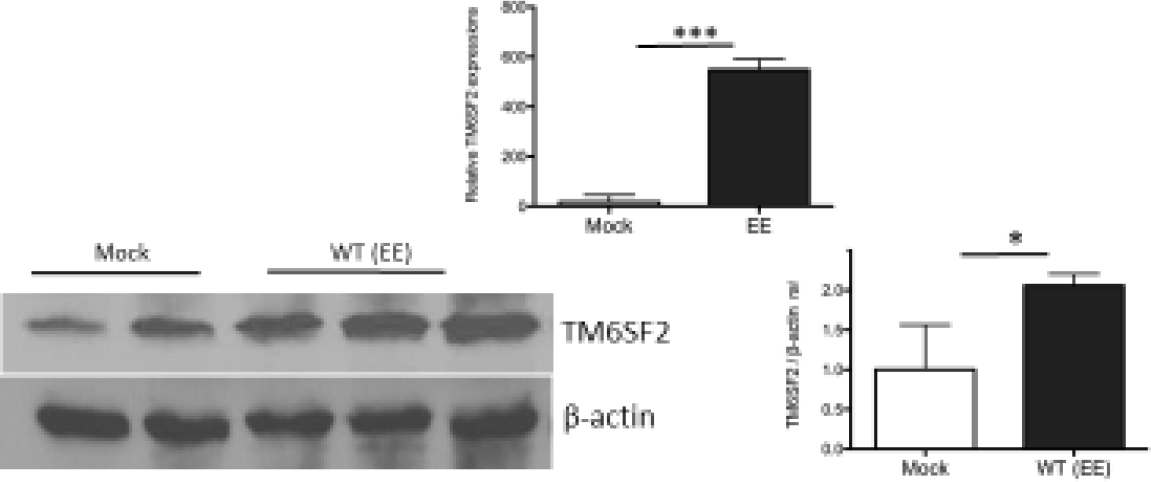
Detection of *TM6SF2* gene overexpression by qRT-PCR and western blotting. Upper: qRT-PCR for measuring *TM6SF2* gene from the cell lines expressing WT *TM6SF2* (EE), and the empty plasmid (Mock); Lower: western blot for detecting WT TM6SF2 levels (EE). Statistical analyses were done using T-Test, *P<0.05, ***p <0 001.

**Figure 2B.**
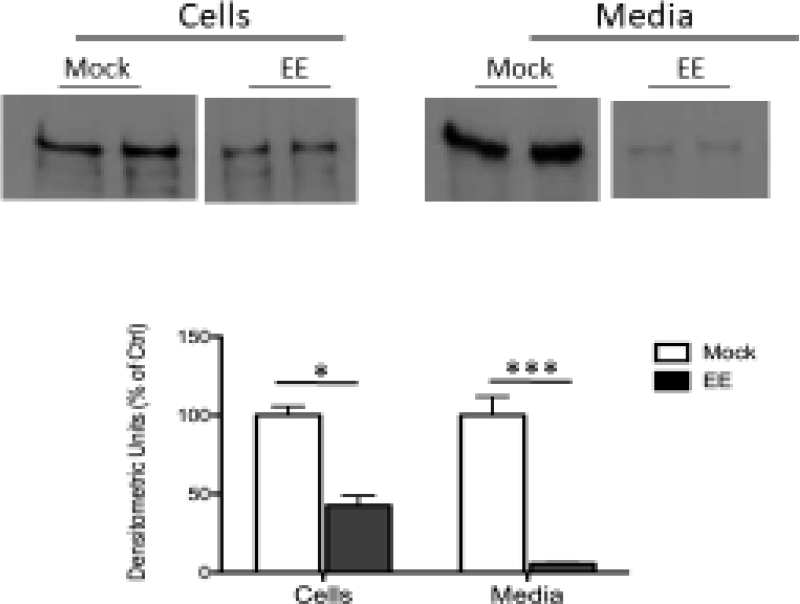
Overexpression of WT *TM6SF2* or Mutant *TM6SF2* (Elb7K) in Hep62 cells both significantly decreased apoB secretion. The experiment protocol was the same as described in Fig 1B. Upper: labeled apoB 100 Lower: Densitometric data from the upper gel Lower Mock: the empty vector; EE: WT *TM6SF2*, The data are expressed the mean ± standard deviation of three Independent experiments. *p<005; ***p <0.001.

**Figure 2C.**
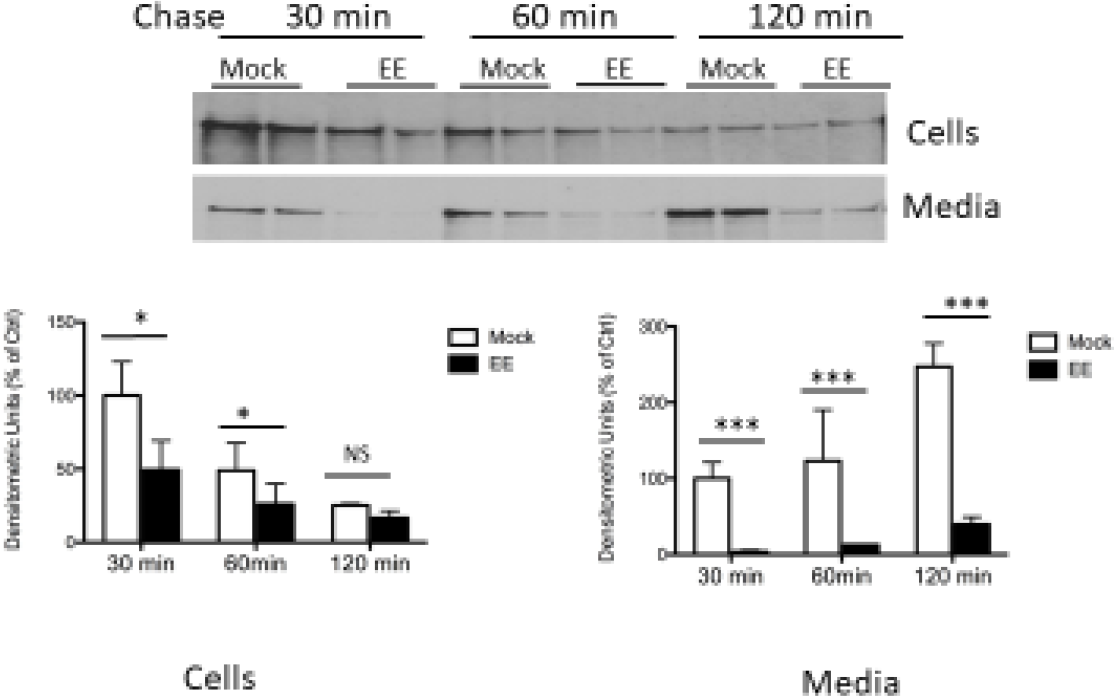
Overexpression of WT *TM6SF2* significantly decreased apoB secretion assessed by pulse chase labeling of apoB Upper: the cells were pulse-labeled for 20 min with [^35^S] Methionine and the medium was changed to one without label and collected at 30 min, 60min or 120min. Lower: Densitometric data from the gel. The data are expressed the mean ± standard deviation of three independent experiments. *p < 005 ***p < 0.001.

### Crispr-induced knockout of Tm6sf2 in McA rat hepatoma cells

**Figure 3A** provides data validating the successful deletion of *TM6SF2*. Two lines of McA cells in which *Tm6sf2* had been deleted (KO 1,2), along with a line having intact, un-edited *Tm6sf2* (Ctrl) were radiolabeled with [35S] methionine for 2 hrs for determination of cell and media newly synthesized, radiolabeled APOB100 and APOB48 (**Figure 3B**). The autoradiogram depicts the APOB100 and APOB48 bands in cells and media (**top**) and these were quantified by densitometry (**bottom**). The **left panel** suggests some increase in APOB100 but not APOB48 in cells of the two KO lines versus the Ctrl line. In contrast, there were clear increases in both APOB100 and APOB48 in the media of the two KO lines versus the Ctrl line (**right panel)**. The response of McA cells to incubation with OA was complex (**Figure 3C**). First, there were increases in newly synthesized cell APOB100 in response to OA in Ctrl but not in KO cells (**far left**). On the other hand, similar to the baseline data for APOB48 depicted in Figure 3C, there was no difference in cell APOB48 between Ctrl and KO cells and, in addition, no increase in APOB48 in response to OA in either Ctrl or KO cells (**second from left**). In the media (**two right panels**), OA increased APOB100 secretion in both the Ctrl and KO cells but, as was the case in cells, the response to OA was reduced in the KO cells, which had greater secretion of APOB100 without OA. The response of APOB48 secretion to OA in both Ctrl and KO cells appeared to be similar to that of APOB100.

**Figure 3A.**
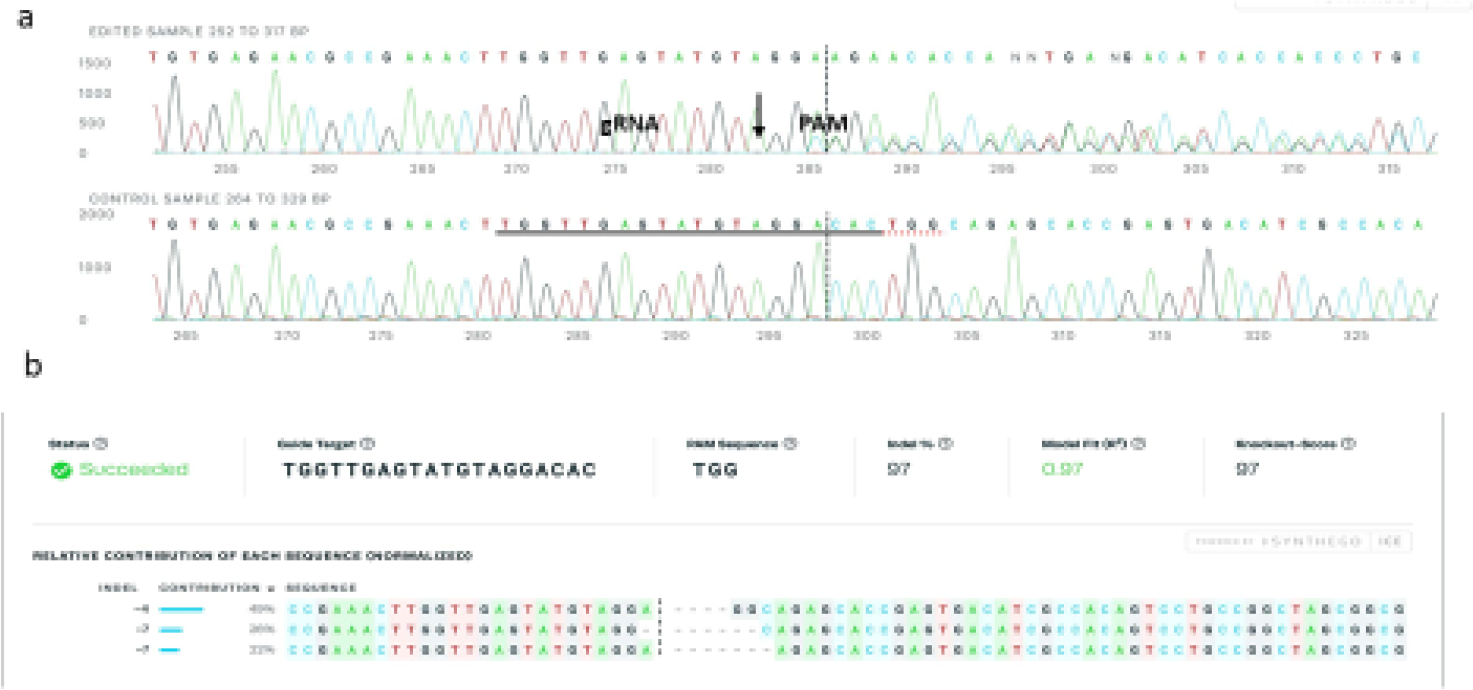
ICE (inference of CRISPR edits) -Coupled Sanger sequencing analysis determines indels of Tm6sf2 gene generated by CRISPR-Cas9. a). The sequencing comparison between the control clone and the edited clone shows the indels are generated spanning the cut site. The alignment window marks the alignment region between the control and target traces, and the inference window marks the region around the predicted sites of genome edition b) The indel plot displays the sizes of indels and their relative prevalence of the indel mixture.

**Figure 3B.**
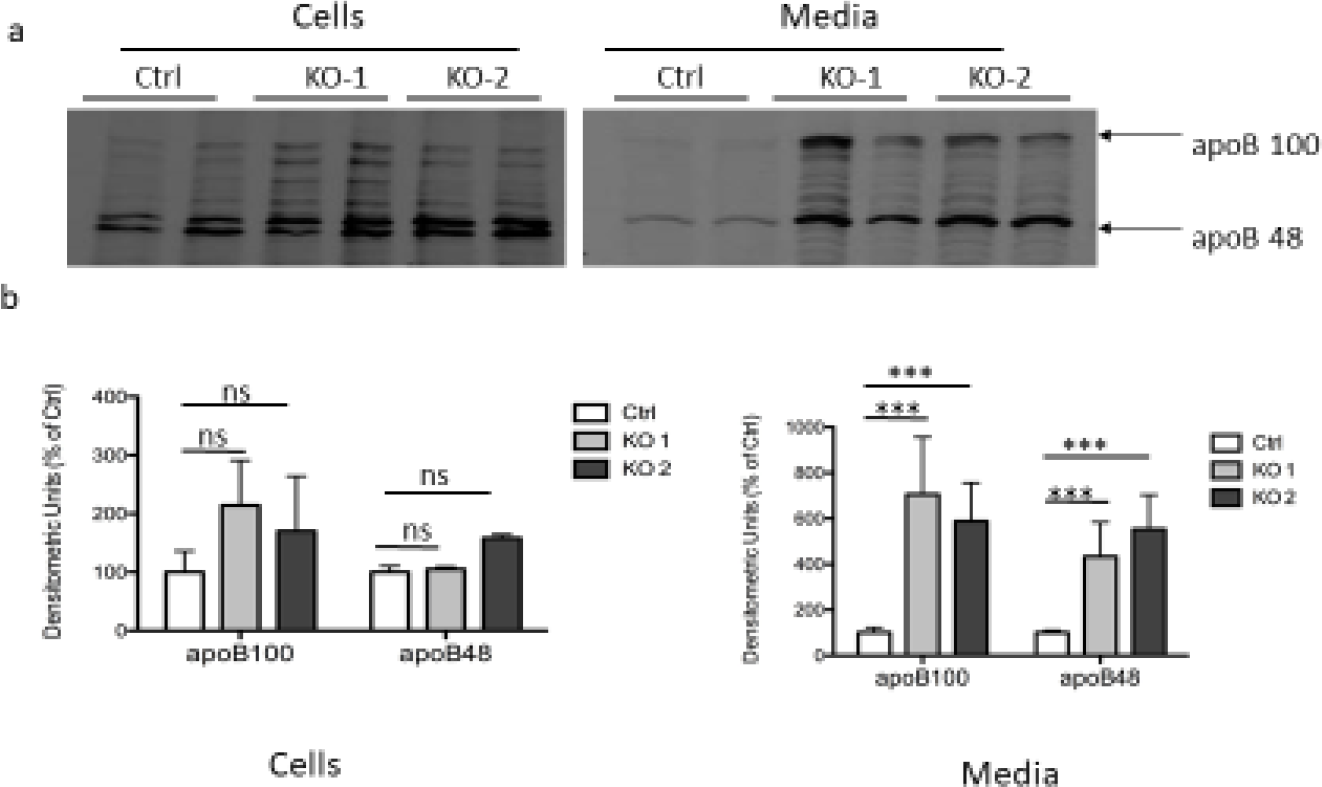
apoB secretions were both significantly increased in two *Tm6sf2* knockout cell lines. The two single cell clones with disrupted *Tm6sf2* reading frame were incubated with and without OA and labeled with [^35^S] methionine for 2 hours The method was described as Fig 1B. Upper panel: apoB labeled with [^35^S] in cells and media. Lower panel: Densitometric data from upper. The data are expressed the mean ± standard deviation of three independent experiments, ***p < 0.001; ns: not significant.

**Figure 3C.**
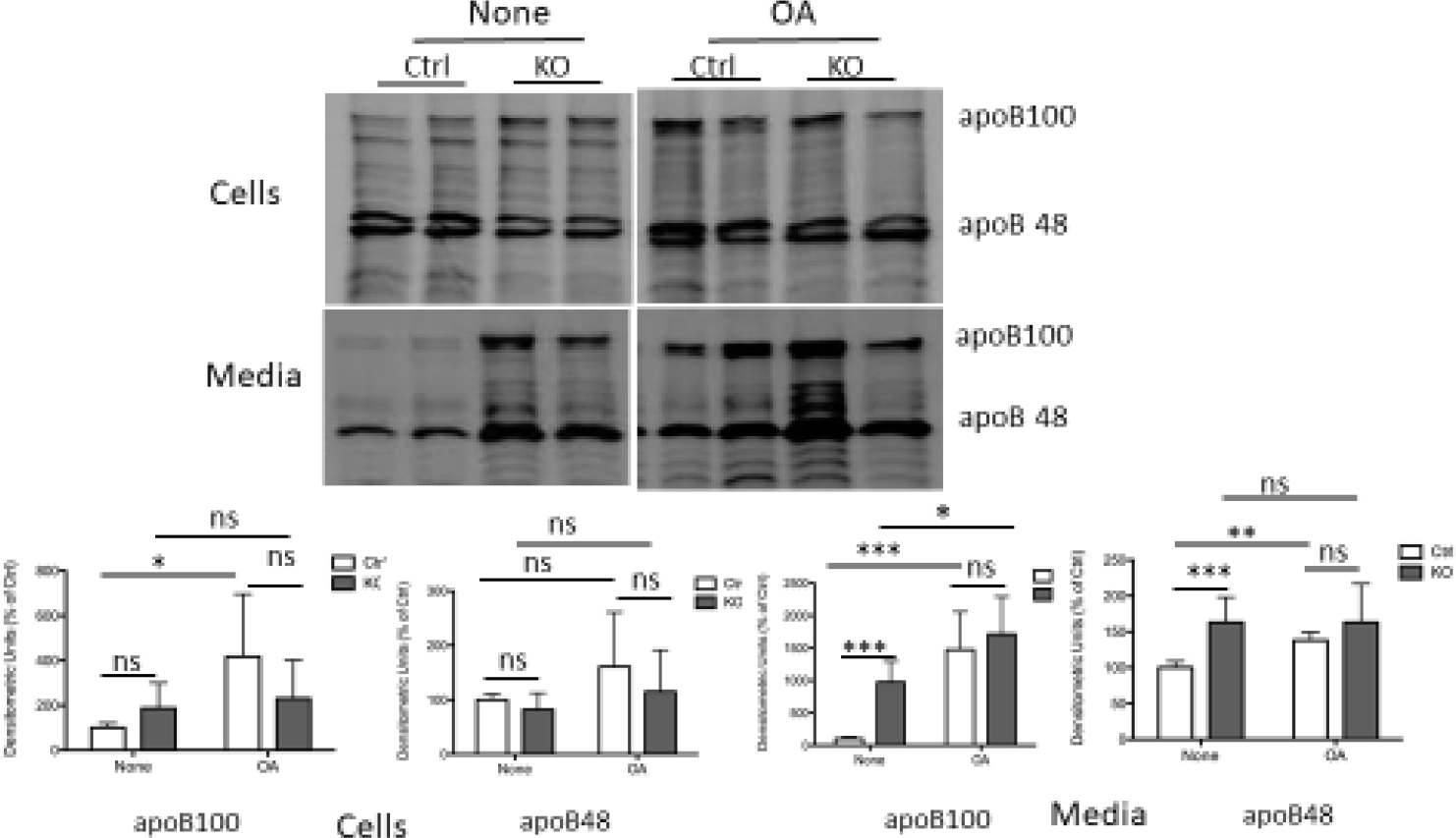
Oleic acid treatment stimulated apoB secretion in WT cells more than in Tm6sf2 KO cells. The cells were incubated with OA for 2h, and then labeled with [^35^S] methionine for 2h with or without OA. Upper panel: SDS PAGE autoradiographs. Lower panel:. Densitometric data from upper. The data are expressed the mean ± standard deviation of three independent experiments. *p < 005, **p < 0.01. ***p < 0 001; ns: not significant

In view of the increases in APOB secretion in the McA KO cells, we examined TG synthesis and secretion using ^14^C OA (**Figure 4A**). After a 4 hr incubation with radiolabeled 0.4 mM OA, ^14^C TG accumulation in the KO cells was double that present in Ctrl cells **(left side)**. In contrast to increased radiolabeled cell TG, ^14^C TG in the media was reduced by almost 50% in the KO cells compared to Ctrl cells (**right side**). The increased radiolabeled ^14^C TG in the KO cells (**Figure 4A**) was paralleled by a 30% increase in TG mass measured enzymatically (**Figure 4B**). Additionally, treatment of KO cells with 0.4 mM OA for 6 hrs results a marked increase in neutral lipid droplets, assessed by BODIPY staining, compared to Ctrl cells (**Figure 4C**). The striking dissociation of APOB secretion and TG secretion led us to examine the size of secreted VLDL by transmission electron microscopy (**Figure 4D, left panel**). There was a clear shift in the secretion of VLDL to particles with smaller diameters from the KO cells compared to the Ctrl cells.

**Figure 4A.**
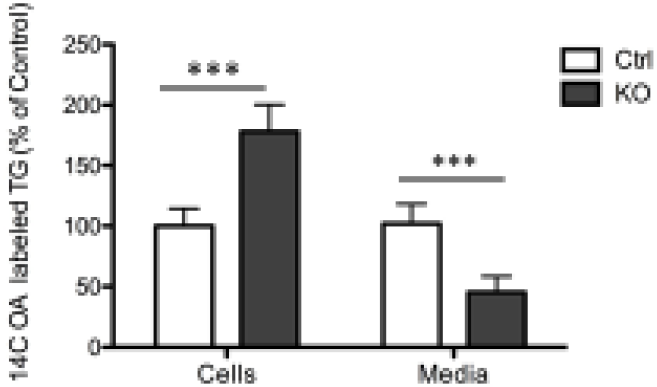
Increased intracellular triglyceride accumulation and decreased triglyceride secretion in Tm6sf2 knockout cells. The cells were labeled with ^14^C OA for 4hrs. Lipid were extracted from cell and medium, and the extracts were separated by thin layer chromatography. TG spots were scraped for ^14^C activity using liquid scintillation counter. The data are expressed the mean ± standard deviation of three Independent experiments. ***p < 0.001

**Figure 4B.**
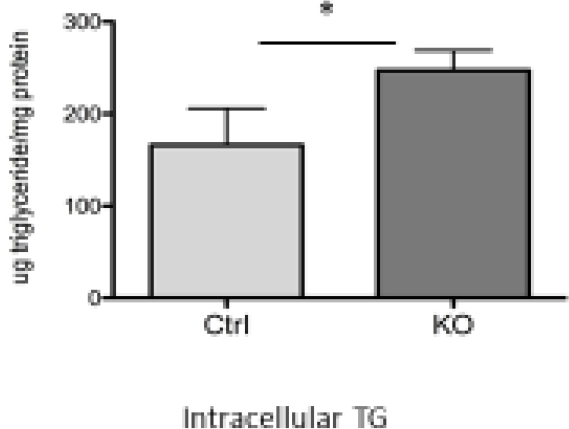
TG content of McA cells was measured using Colorimetric triglyceride assay. The cells were treated with 0.2mM OA for 16 h, and harvested and suspended in 5%NP-40 solution. Total cellular triglycerides were quantified using an enzymatic triglyceride method. The total cellular triglyceride content was normalized by total cellular protein. Each bar represents average TG content from the cells of 3 T75 flasks, *p<0.05.

**Figure 4C.**
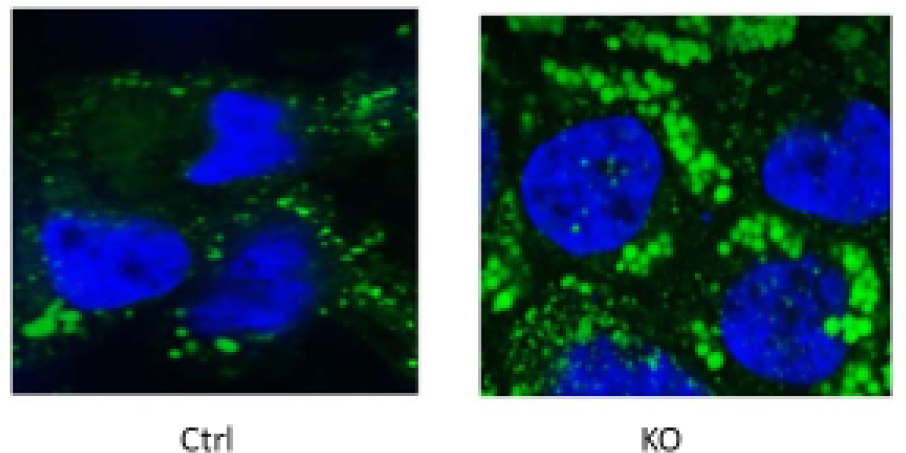
Lipid droplets were stained with BODIPY. The cells were treated with 0.4mM OA for 6h. Then intracellular lipid droplets were stained with the BODIPY (493/503, green neutral lipid stain). The nucleus was stained with DAPI.

**Figure 4D.**
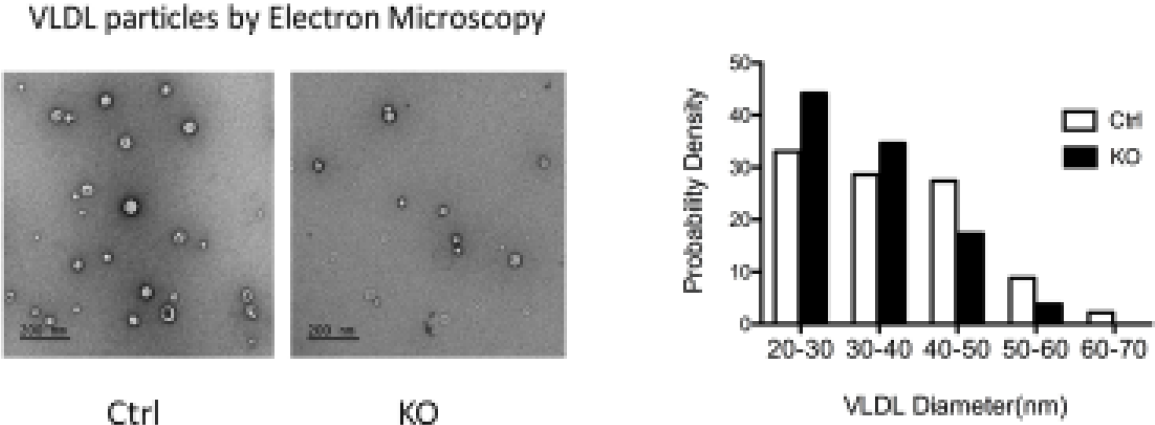
*Tm6sf2* deficient cells secreted smaller VLDL particles than WT cells. Cells were treated with OA for 16h and the media collected for sucrose density gradient ultracentrifugation for 65h. 1ml of VLDL on the top of tube was collected and 5 µl of fixed suspension was added onto glow discharged carbon coated 400 mesh Cu/Rh grid stained with 1% aqueous uranyl acetate. Stained grids were Imaged using transmission electron microscope (EM). Left: VLDL particle from EM micrograph. Right: The size distribution of VLDL particles analyzed by Image J.

RNA seq was performed as described in Methods. A heatmap of genes related to endoplasmic reticulum (ER) stress response was constructed with data of from *Tm6sf2* KO versus WT cells (**Figure 5A**). TM6SF2 deficiency resulted in significant downregulation of several genes (**left panel**) encoding proteins (**right panel**) involved in the unfolded protein response (UPR) to ER stress (the proteins are shown for ease of reading). The genes included IRE-1a, PERK, ATF6, eIF2a, and CHOP. Expression of the gene for XBP1, a transcription factor for secretory proteins was also reduced. To determine if these changes in gene expression in the KO cells were reflected in protein content, we carried out Western blotting studies (**Figure 5B**). There were clear reductions in p-eIF2α, IRE1α, and CHOP protein levels, and a modest decrease in BIP.

**Figure 5A.**
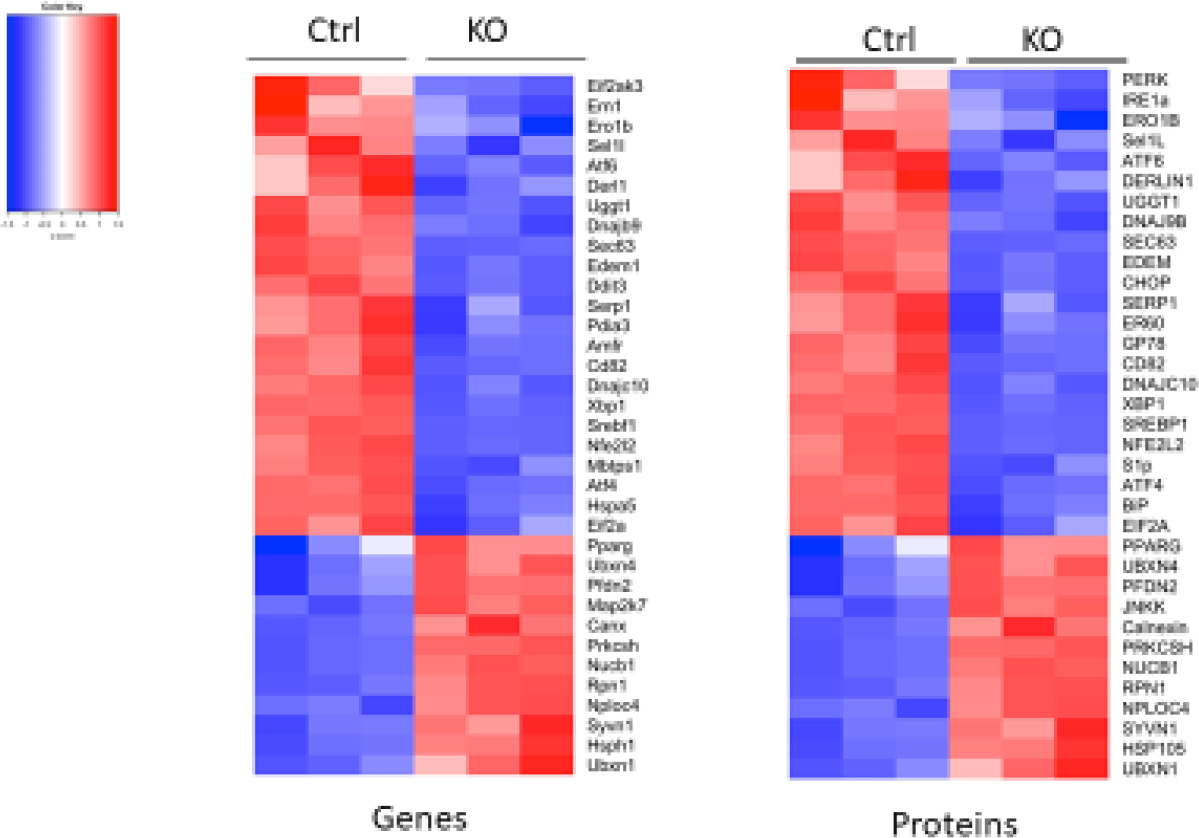
Heatmap of genes related to endoplasmic reticulum (ER) stress response from RNA-Seq data of *Tm6sf2* KO cells vs Ctrl cells. TM6SF2 deficiency leads to reduced expression of genes for three major ER proteins involved in the unfolded protein response(UPR), including IRE-la, PERK, and ATF6 as well as the gene for the transcription factor XBP1 and the gene for CHOP. The heatmap color range is from red for positive Z-scores values to blue for Z-score negative values. Genes are on the left and proteins (provided for ease of reading) are on the right.

**Figure 5B.**
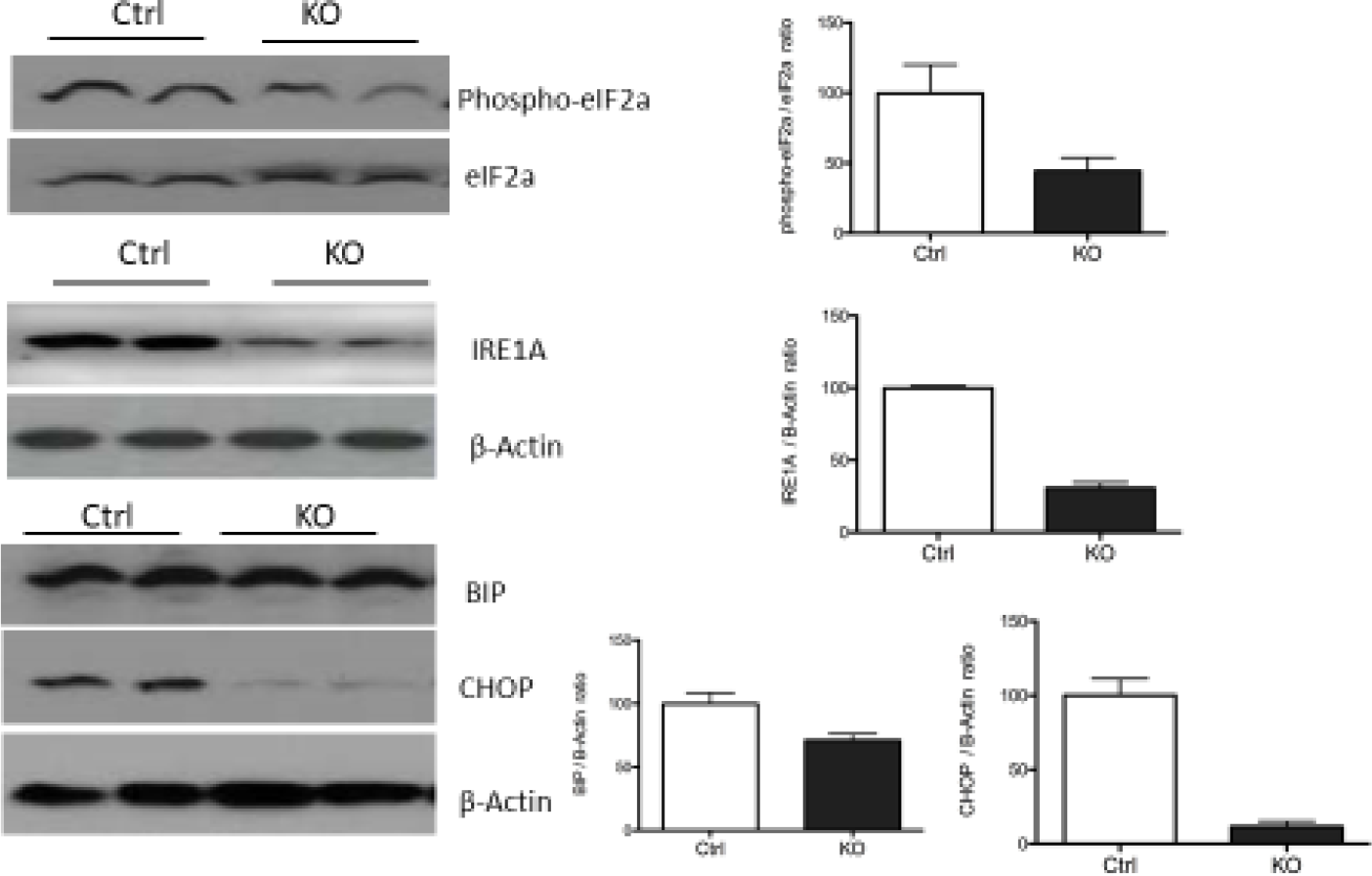
Western blots Tm6sf6 KO and Ctrl cells for ER stress markers p-elF2, BIP, IRE 1α, and CHOP, β-Actin and elF2α are loading controls

## Discussion

Between 2008 and 2013, 6 GWAS studies identified variants in a region on chromosome 19 associated with plasma TG and cholesterol levels ^9, 22–25^ as well as NAFLD ^9, 26^. This locus, termed 19p12 or NCAN-TM6SF2-SUGP1-CILP2, contains more than a dozen genes, none of which had previously been implicated in lipid metabolism. In 2014, Kozlitina et al. identified a missense loss-of-function variant, p.E167K in TM6SF2 that was associated with NAFLD ^10^. Those investigators knocked-down *Tm6sf2* in wild-type mice with a shRNA and observed significant hepatic steatosis and decreased TG secretion *in vivo* ^10^. Additionally, they found that the TM6SF2 variant encoding p.E167K was associated with low plasma levels of TG and LDL-C, but not HDL-C, in the DHS, Dallas Biobank. At the same time, Holmon et al. identified TM6SF2 as a variant associated with lower plasma lipids ^11^. A few months later, Mahdessian *et al.* reported a significant, positive relationship between hepatic expression of WT TM6SF2 and levels of plasma TG, suggesting that TM6SF2 is the putative functional gene in the 19p12 locus responsible for the observed relationships with plasma TG levels ^27^.

The studies above increased interest in this gene and its protein and that led to publications providing several clear and important insights regarding the physiologic consequences of the p.E167K variant. First, it is clear that the variant is tightly associated and probably causal for NAFLD ^28, 29^. Second, in populations, the variant is associated with modest to moderate reductions in plasma lipid levels ^10, 11, 28^. Third, the p.E167K variant results in decreased secretion of TG with either no change or an increase in APOB100 and APOB48 secretion in mice ^10, 11, 30^. The latter is consistent with demonstration that small VLDL are being secreted, likely due to reduced incorporation of TG into each VLDL particle. What is less clear is how TM6SF2 is linked to VLDL lipidation and why there is a dissociation of the rates of TG and VLDL APOB secretion in the variant. The exact molecular mechanism underlying the role of TM6SF2 in lipidation of nascent VLDL (second-step lipidation) remains to be determined, although a physical interaction between TM6SF2 and APOB, possibly enhanced by the presence of ERLINS 1 and 2 could enable that process ^31^.

The dissociation of the rates of secretion of VLDL TG and APOB in TM6SF2 knockout mice is, at first glance, in contrast to studies in mice where diet induced obesity results in increased secretion of both TG and APOB ^32^ and in humans with insulin resistance, diabetes, or metabolic syndrome ^33^. However, there are several examples in human tracer kinetic studies where dissociation of TG and APOB secretion have been observed, including a high carbohydrate diet, where TG secretion increased without a change in APOB secretion ^34^ and weight loss in obese individuals with metabolic syndrome where TG secretion decreased more than APOB secretion ^35^. In the first published human tracer study comparing people homozygous for the p.E167K variant and people without the variant, Boren et al observed similar and very significant reductions in the rates of secretion of both TG and APOB100, which fell approximately 35 and 45%, in the large, TG-enriched VLDL1 fraction ^14^.

Our results have some similarities and some differences from those described above. Most striking is our demonstration of a significant increase of 38% in the secretion of VLDL APOB100 with no change in VLDL TG production in sib-pairs with or without the p.E167K variant for TMSF2. Thus in sharp contrast to the data from Boren et al.^14^ where the data suggest that the p.E167K variant resulted in secretion of fewer VLDL particles of the same size as seen in people without the variant, our results indicate increased secretion of smaller particles, a conclusion supported by the lower ratio of TG PR to APOB100 PR in the siblings with the variant. There are several possibilities for these differences, including different subject cohorts (Lancaster Amish versus Scandinavians), very different levels of plasma TG, LDL, and APOB100 in the Amish without p.E167K compared to their Scandinavian counterparts, and the protocol used for the kinetic tracer studies. Boren et al. fed a large high fat meal to the subjects at the beginning of the study and isolated VLDL in two fraction; we fed very low fat meals every 2 hours throughout the 24 hour study period and examined the entire VLDL fraction.

We were not expecting to see a significant increase in VLDL APOB100 secretion, despite studies in mice where some increases in APOB secretion had been observed. We turned, therefore, to two immortalized cell models of APOB secretion, HepG2 and McA cells. The results in HepG2 cells supported a modest increase in APOB100 when TM6SF2 was knocked-down by a specific shRNA; this finding was consistent with some, but not all of the prior cell-based studies. Of interest was the significant reduction in APOB100 secretion when TM6SF2 was overexpressed in HepG2 cells, a finding consistent with that of Ehrhardt et al. in human hepatocytes with overexpression of TM6SF2 ^37^. In our McA cells, Crispr-based deletion of *Tm6sf2* was associated with clear increases in the secretion of both APOB100 and APOB48 that was accompanied by increased accumulation of newly synthesized radiolabeled TG and increased TG mass in the cells, decreased secretion of newly synthesized TG into the media, increased lipid droplets in McA knockout cells upon loading with OA, and secretion of smaller VLDL particles. Of particular interest, RNA-Seq data indicates reduced ER stress in the McA knockout cells, results supported by Western blotting studies. We had previously demonstrated that fatty acid-induced ER stress inhibited APOB secretion in McA cells and mice ^38, 39^.

Integrating our results into the large prior literature is a daunting task and certainly made more difficult by the heterogeneous non-human data and of course, the stark difference between the two human kinetic studies. With caution, therefore, we suggest the following model for humans: TM6SF2 interacts physically with APOB100 allowing addition of ER synthesized TG to nascent, relatively lipid poor VLDL. In the presence of normal levels of TM6SF2, this interaction leads to a fully lipidated “mature VLDL” that moves to the Golgi and then to the plasma membrane for secretion. If the mass of TM6SF2 is reduced, as seems to be the case for the p.E167K variant^10^, some VLDL can still move forward toward secretion, with less TG on each VLDL particle (this may suggest that the interaction of nascent VLDL with TM6SF2 is iterative), but other VLDL do not mature and, possibly due to incomplete folding of APOB around the VLDL particle, induce ER stress and are targeted for degradation. Data from rodents and humans have long suggested that hepatocytes constitutively synthesize APOB and secrete as many VLDL as are needed to maintain hepatic lipid homeostasis ^40^, and TM6SF2 may act to either add lipid to nascent VLDL or, when additional lipid is not needed, or cannot be integrated into nascent VLDL, target some of those particles for degradation, possibly by what has been termed post-ER presecretory proteolysis (PERPP) ^41^.

Finally, how would this model allow for increased secretion of APOB? We postulate that when hepatic TG levels are in excess, insufficient TM6SF2 will likely lead to TG or APOB mediated ER stress and greater degradation of nascent or incompletely lipidated VLDL via PERPP, whereas when hepatic lipids are normal or low, reduced or absent TM6SF2 will be associated with increased secretion of poorly lipidated VLDL. Clearly, there is more work to do if we are to progress toward a more complete understanding of the complex role of TM6SF2 in hepatic lipid metabolism and both the secretion of VLDL and the risk of NALFD associated with the p.E167K variant.

## Data Availability

All data produced in the present study are available upon reasonable request to the authors.

## Acknowledgements

We are grateful for the efforts and support of the Amish Research Clinic nurses, technicians, and staff in Lancaster, PA. This study would not have been possible without the outstanding support of the Amish research participants. We also thank the Irving Institute for Clinical and Translational Research Bionutrition Unit for developing and providing all study meals.

